# Prevalence and seroprevalence of COVID-19 infection among older people: A scoping review based on population-based studies in 2020-2022

**DOI:** 10.1101/2024.03.08.24303942

**Authors:** Jingxin Lei, Phyumar Soe

**Author notes:** Corresponding author at: School of Population and Public Health, University of British Columbia, BC V6T 1Z4, Canada.

## Abstract

**Background:** Accurate estimates of the prevalence of infections play an important role in COVID-19 surveillance. Older people are known to have higher risks of severe outcomes after infection, but whether they also have a higher infection rate remains unclear. To obtain estimates of COVID-19 prevalence among older people, we synthesized evidence from RT-PCR-based prevalence and serological studies.

**Methods:** We conducted a scoping review using a comprehensive search of MEDLINE (Ovid), Embase (Ovid), Europe PMC, ClinicalTrials.gov, and the WHO COVID-19 Research Database from December 2019 to Oct 2022. We included population-based cross-sectional (sero)prevalence studies among older people (i.e., people aged >= 65 +/-5 years) who were tested for SARS-CoV-2 infection using RT-PCR tests, antigen tests, or serological tests. Studies that were conducted solely in institutional housing were excluded. Eligible studies were extracted and critically appraised. We described and mapped the prevalence (tested by RT-PCR or antigen tests) and seroprevalence (tested by serological tests) by geographical area and time. We then compared the estimated prevalence with WHO-reported prevalence and the prevalence among younger age groups from the same study.

**Results:** We identified 202 (sero)prevalence estimates from 126 studies, covering 50 countries up to October 2022. Of the 126 studies, 28 studies estimated RT-PCR-based prevalence; 104 studies estimated seroprevalence, ranging from 0% in Jordan to 22.5% in the United States in 2020, from 0.41% in Brazil to 98% in Chile in 2021. In the year 2020, prevalence of COVID-19 ranged from 0.0006% in China, to 52.8% in Brazil, while in 2021, prevalence ranged from 0.06% in England to 41.1% in Brazil. The ratio of the reported prevalence to estimated prevalence ranged from <0.01 to 77.50, where 86% (24/28) studies estimated a higher prevalence than WHO reported and half of them estimated >10 times higher prevalence. One third of studies (32%, 9/28) estimated a higher prevalence in older people compared with younger people.

**Conclusions:** Our findings suggest that underreporting of COVID-19 cases among older people may exist extensively worldwide. Compared with younger groups, older people were less likely to be infected with COVID-19 in two thirds of the studies through the first two years of the pandemic.

## Background

The COVID-19 pandemic is one of the most distributed pandemics in human history^1^. As of March 10, 2023, 676,609,955 cases and 6,881,955 deaths have been reported^2^. However, the reported cases may not capture the actual burden of the pandemic, partially due to limited test availability and accessibility^3,4^, asymptomatic infections^5^, and the extensive allocation of self-test kits^6^. For instance, according to a global survey covering respondents from Africa, Americas, Eastern Mediterranean, Europe, Southeast Asia, and Western Pacific, self-testing were available in 101 out of 139 countries in early 2022^7^.

COVID-19 seroprevalence and prevalence studies across the world have described the spread of the virus over time and in different regions, age-groups and socioeconomic profiles. Prevalence studies estimate prevalence of SARS-CoV-2 virus RNA (based on nucleic acid amplification tests, or NAAT in short) or antigen of SARS-CoV-2 (antigen based). Reverse transcription polymerase chain reaction (RT-PCR) is the most widely used method among NAATs. It is one of the most accurate tests of active infections but more expensive and time-consuming than antigen tests, which are more accessible alternatives but less accurate. Seroprevalence studies estimate prevalence of SARS-CoV-2 antibodies as a result of vaccination or infection, depending on the types of antibodies. Antibody remains detectable months after infection or vaccination^8^. Compared with RT-PCR tests, both antibody and antigen tests can be conducted quicker and cheaper, thus are more commonly used. Because of their ability to detect past infection or vaccination weeks or months ago, antibody tests have been used to estimate COVID-19 prevalence, as well as to estimate the extent of case underreporting^9,10^. However, based on WHO’s definition^11^, a confirmed case is either an individual with a positive NAAT result or a suspect case, based on clinical and/or epidemiological criteria, with a positive antigen result. Literature suggested that antibody tests tend to provide a higher positivity rate than PCR tests^12^. Using a seroprevalence-based estimate of prevalence may lead to a higher estimate of COVID-19 infections. No review has analyzed PCR-based prevalence studies at the initiation of this study.

Older people are among the most vulnerable populations to experience severe outcomes after COVID-19 infection^13–15^. However, whether older people are also more vulnerable to COVID-19 infection itself remains unclear. Since a recent review from Bergeri et al has analyzed seroprevalence studies comprehensively^10^, in this review we aim to 1) describe COVID-19 seroprevalence and prevalence among older people across region and time, 2) assess the extent of underestimation of COVID-19 infections based on prevalence studies, and 3) compare the prevalence between older people and younger groups.

## Methods

### Information sources and search strategy

We searched the following electronic databases: MEDLINE (Ovid), Embase (Ovid), and Europe PMC from December 2019 to Oct 19, 2022. Embase and Europe PMC were searched to include preprints. We also searched ClinicalTrials.gov and the WHO COVID-19 Research Database to include COVID-19 prevalence studies registered in these databases. Backward and forward citation searching on included studies was conducted. The comprehensive search strategies with MeSH terms are summarized in Table 1. For database where MeSH terms were not available, following keywords are searched: (“COVID-19” or “SARS-CoV-2” or “coronavirus”) and (“prevalence” or “incidence” or “seroprevalence”) and (“covid-19 testing” or “nucleic acid test” or “antigen test” or “rapid test” or “antibody” or “PCR” or “RT-PCR” or “polymerase chain reaction”) and (“older people” or “older adult” or “aged people”). The review protocol^16^ is registered on Open Science Framework^17^(Registration DOI: doi.org/10.17605/OSF.IO/37RZS).

**Table 1.** Comprehensive search strategy.

|  |  | MEDLINE<sup>1</sup> | Embase |
| --- | --- | --- | --- |
| 1 | COVID-19/ | 192270 | 121814 |
| 2 | SARS-CoV-2/ | 139231 | 38177 |
| 3 | (covid* or coronavirus or corona virus or sars-cov-2 or pandemic).mp.<br>[mp=title, book title, abstract, original title, name of substance word,<br>subject heading word, floating sub-heading word, keyword heading word,<br>organism supplementary concept word, protocol supplementary concept<br>word, rare disease supplementary concept word, unique identifier,<br>synonyms] | 348147 | 419612 |
| 4 | 1 or 2 or 3 | 348147 | 419612 |
| 5 | prevalence/ | 335697 | 879533 |
| 6 | incidence/ | 296161 | 525645 |
| 7 | seroprevalence.mp. or Seroepidemiologic Studies/ | 35437 | 39569 |
| 8 | prevalence.mp. | 840829 | 1290274 |
| 9 | 5 or 6 or 7 or 8 [prevalence/seroprevalence] | 1111206 | 1755607 |
| 10 | covid-19 testing/ or covid-19 nucleic acid testing/ | 9030 | 6920 |
| 11 | (nucleic acid test* or antigen test* or rapid test*).mp. | 16272 | 33779 |
| 12 | (estimat* or underreport*).mp. | 1371317 | 1745061 |
| 13 | 10 or 11 or 12 [testing] | 1392591 | 1780575 |
| 14 | exp aged/ or exp geriatrics/ or exp geriatric psychiatry/ or exp geriatric<br>nursing/ or exp *geriatric psychiatry/ or exp *dental care for aged/ or exp<br>*health services for the aged/ or (elder* or eldest or frail* or geriatri* or<br>old age* or oldest old* or senior* or senium or very old* or<br>septuagenarian* or octagenarian* or octogenarian* or nonagenarian* or<br>centarian* or centenarian* or supercentenarian* or older people or older<br>subject* or older patient* or older age* or older adult* or older man or<br>older men or older male* or older woman or older women or older<br>female* or older population* or older person*).ti,ab,kf. | 3668977 | 3904096 |
| 15 | cross-sectional studies.mp. | 451139 | 18593 |
| 16 | mass screening/ or anonymous testing/ | 115098 | 60437 |
| 17 | 9 or 15 or 16 [upated prevalence/seroprevalence] | 1518900 | 1817065 |
| 18 | Serologic Tests/ or Serologic Tests.mp. | 24427 | 88223 |
| 19 | Serology.mp. or Serology/ | 29341 | 106912 |
| 20 | immunoassay/ or fluoroimmunoassay/ or immunoenzyme techniques/ or<br>immunologic tests/ | 108361 | 4572948 |
| 21 | Antibodies/ or antibod*.mp. | 1256738 | 1600218 |
| 22 | 18 or 19 or 20 or 21 [serologic testing] | 1343971 | 5143721 |
| 23 | polymerase chain reaction/ or real-time polymerase chain reaction/ or<br>reverse transcriptase polymerase chain reaction/ or pcr.mp. or polymerase<br>chain reaction.mp. | 930022 | 1425171 |
| 24 | 13 or 23 [nucleic acid testing] | 2282195 | 3147498 |
| 25 | 22 or 24 [all testing methods] | 3495859 | 7567506 |
| 26 | 4 and 17 and 25 | 10592 | 20739 |
| 27 | 14 and 26 | 2224 | 4451 |
| 28 | limit 27 to (english language and yr="2019 -Current") | 1820 | 3985 |

Note: 1. MEDLINE version: Ovid MEDLINE(R) and Epub Ahead of Print, In-Process & Other Non-
Indexed Citations, Daily and Versions(R) 1946 to September 04, 2020

### Eligibility criteria

This review included cross-sectional or repeated cross-sectional studies using molecular tests, antigen tests, or serological tests (i.e., antibody tests) to estimate the prevalence and seroprevalence of COVID-19 of people in municipalities, regions, states, or countries around the world. Studies were excluded if the study:

i. was not a cross-sectional study,
ii. did not report prevalence or seroprevalence specifically among the older population,
iii. restricted to a specific group of participants only (e.g., people living in residential facilities such as long-term care facilities or nursing homes, or having certain underlying condition such as cardiovascular diseases), and
iv. had a sample size less than 100

For the purpose of our review, older populations were defined as people with a minimum age of 65 ± 5 years. Studies using convenience sampling were also included to gather information as thorough as possible. Only literature with a title and abstract written in English were included. Because of the rapidly evolving state of the COVID-19 pandemic, non-peer-reviewed articles such as pre-prints and scientific reports were included. Conference proceedings alone were excluded.

### Study selection and data extraction

All reference management were performed in Covidence^18^. After the removal of duplicates, titles and abstracts of all search results were assessed for inclusion against our eligibility criteria. Prior to the screening of title and abstract, the screening form was calibrated through pilot testing^19^ with a random sample of 25 records from the literature search by two independent reviewers. At the first stage, two reviewers JL and PS screened all titles and abstracts independently to decide if the record was potentially eligible, irrelevant, or of indeterminate relevance. All potentially eligible records and those of indeterminate relevance were retrieved and screened at the full-text level. Discrepancies were resolved by discussion and consensus between the two reviewers or by a third-party decision. Information of the included studies was extracted according to a pre-specified data extraction form (Table 2). Prevalence of younger age group from the same study was also extracted if available. We did not extract vaccination status as most of the studies didn’t report it. Two reviewers extracted data independently and consensus was reached afterwards.

**Table 2.**
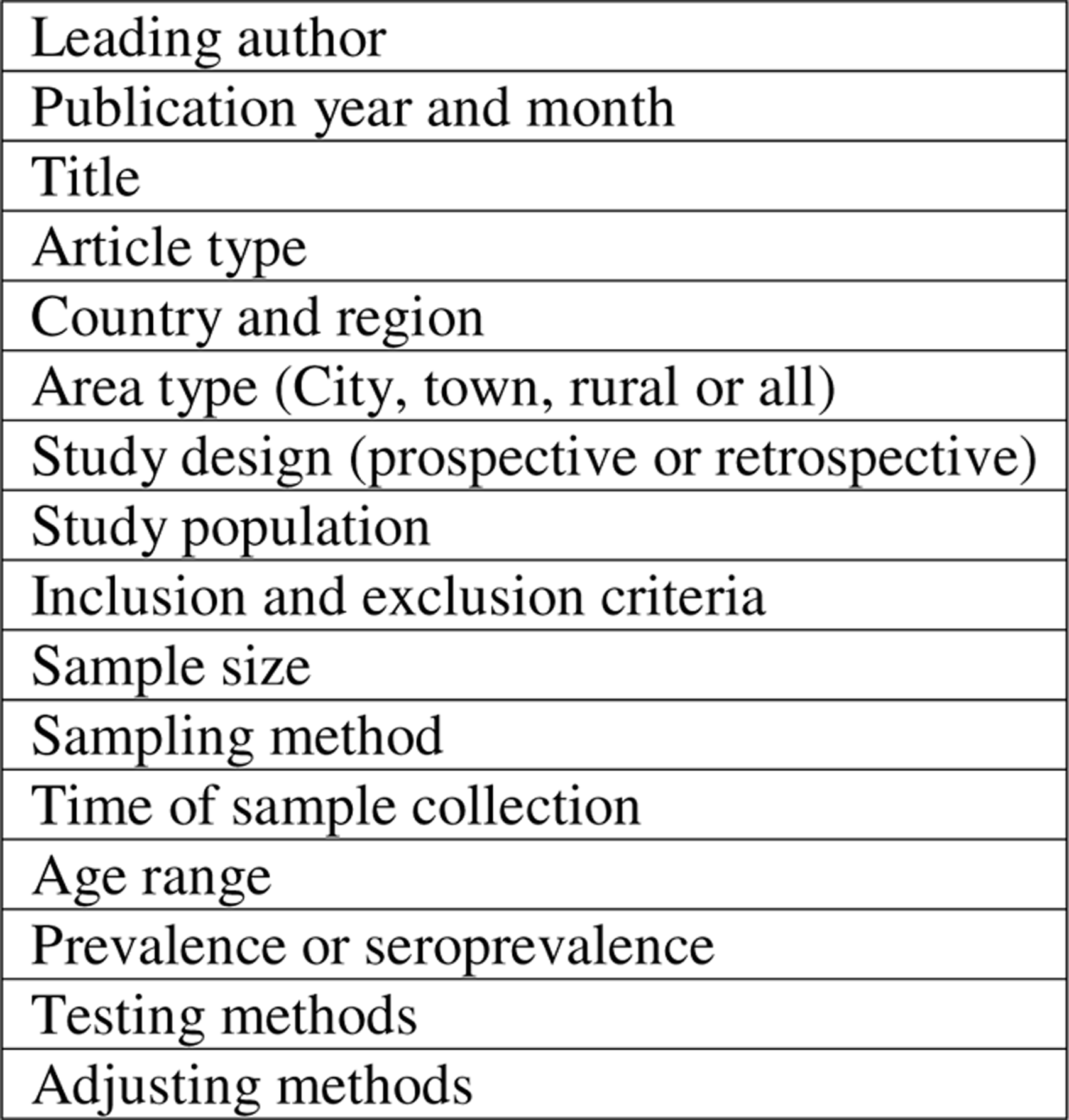
Data extraction form.

### Statistical analyses

A narrative description of the prevalence and seroprevalence studies was presented by geographical region and time. For prevalence studies, we compared the prevalence of COVID-19 based on included studies with reported prevalence in the same country. The reported prevalence was calculated by the reported cases among people aged 65 and above during the sample collection period reported by WHO^23^, divided by total population size aged 65 and above in 2021, reported by United Nations, World Population Prospects^24^. If age-specific cases were not available in a country, we would use all-age prevalence at the same time as a comparator. Analyses were conducted with R language^25^

## Results

### Eligible evaluations

6,248 records were identified in the databases after the comprehensive search as of October 19, 2022. After automatically removing the duplicates by Covidence, 5,163 records were screened. 4,958 records were excluded in the title and abstract screening; and 205 records retrieved for full text. Fifty-nine articles were excluded in the full-text screening due to off-target population (n=29), off-target measurement (n=15), inadequate sample size (n=8), or duplicates (n=7). In total, 126 unique studies were retrieved in 146 records (Figure 1). In these 126 studies, 202 estimates of (sero-)prevalence were extracted. The information extracted for the eligible studies is summarized in Supplementary Table.

**Figure 1.**
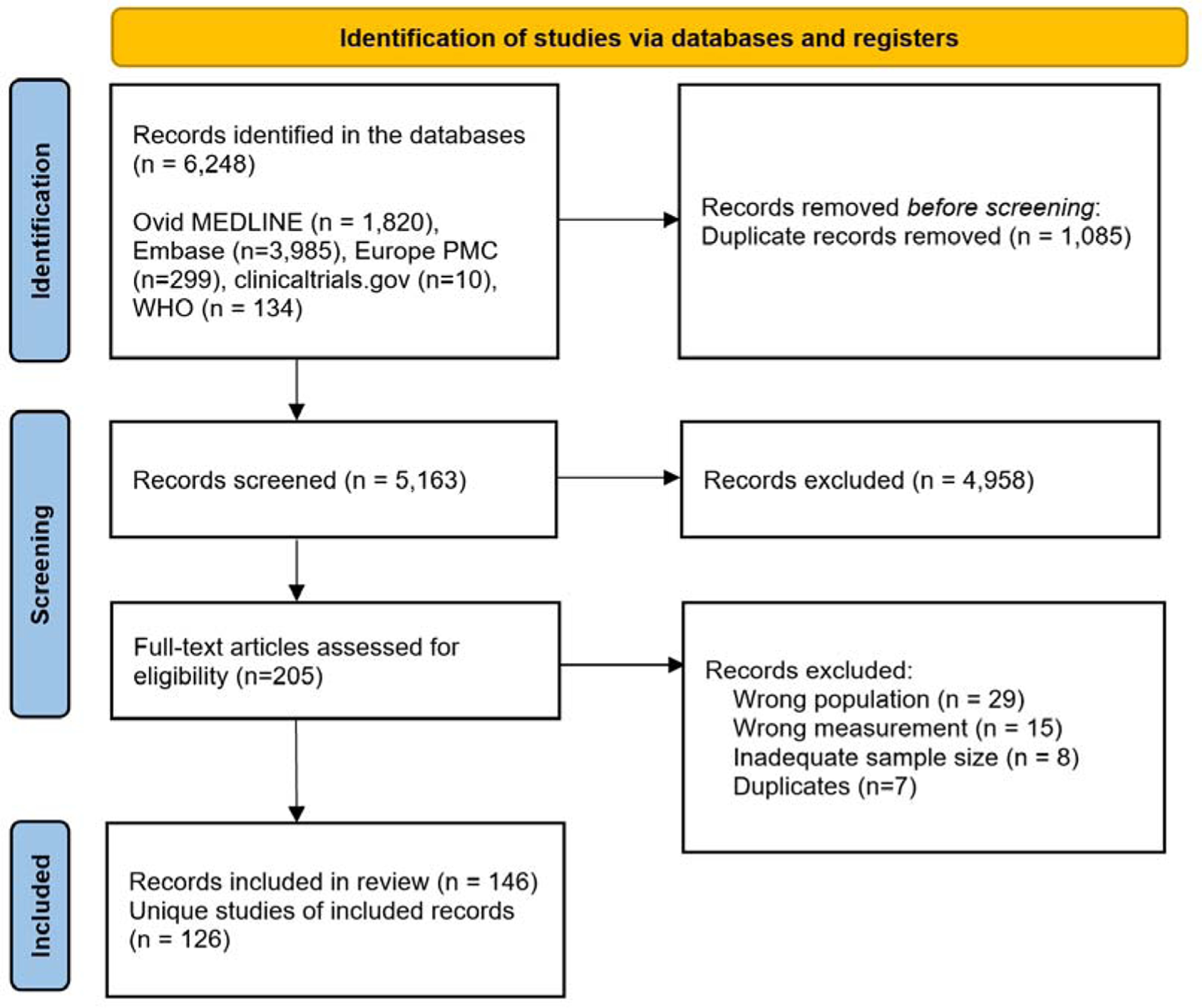
PRISMA Flow Diagram.

### General description of the studies

Of the 126 included studies, samples of 85 studies were collected in 2020, 11 were collected in 2021, 1 was collected in 2022, and 29 were collected in multiple rounds across 2020-2022. The prevalence and seroprevalence estimates over time are shown in Figure 2. Five studies were preprints or scientific reports, while the majority (n=121) of studies were peer-reviewed journal articles. Regions with the most studies were Europe (n=48), North America (n=30), and Asia (n=28). Only 7 studies were conducted in Africa and 13 studies in South America (Figure 3 and 4). No identified studies were conducted in Oceania. Countries with most studies are the United States (26 unique studies with 39 estimates), India (11 unique studies with 19 estimates), and Brazil (8 unique studies with 13 estimates). Ninety studies were prospective cross-sectional studies, 34 studies were retrospective cross-sectional studies, and 2 studies collected seroprevalence prospectively whereas the reported PCR-based prevalence data was retrieved retrospectively from other sources. Half (n=66, 52%) of the studies were randomly sampled from the general population. For non-randomly sampled studies, 15 studies were sampled from the general population, 14 studies were based on samples for COVID-19 testing, and 26 studies were based on samples for other reasons (e.g., routine laboratory tests). Three studies had a different sample scheme for prevalence and seroprevalence^26–28^, and one study reported in three publications sampling from the general population randomly in Russia^29^ but non-randomly in the Kyrgyz Republic^30^ and the Republic of Belarus^31^. The age ranges for older participants were defined as 60 years and above (n=69 studies), 65 and above (n=46 studies), and 70 and above (n=7 studies).

**Figure 2.**
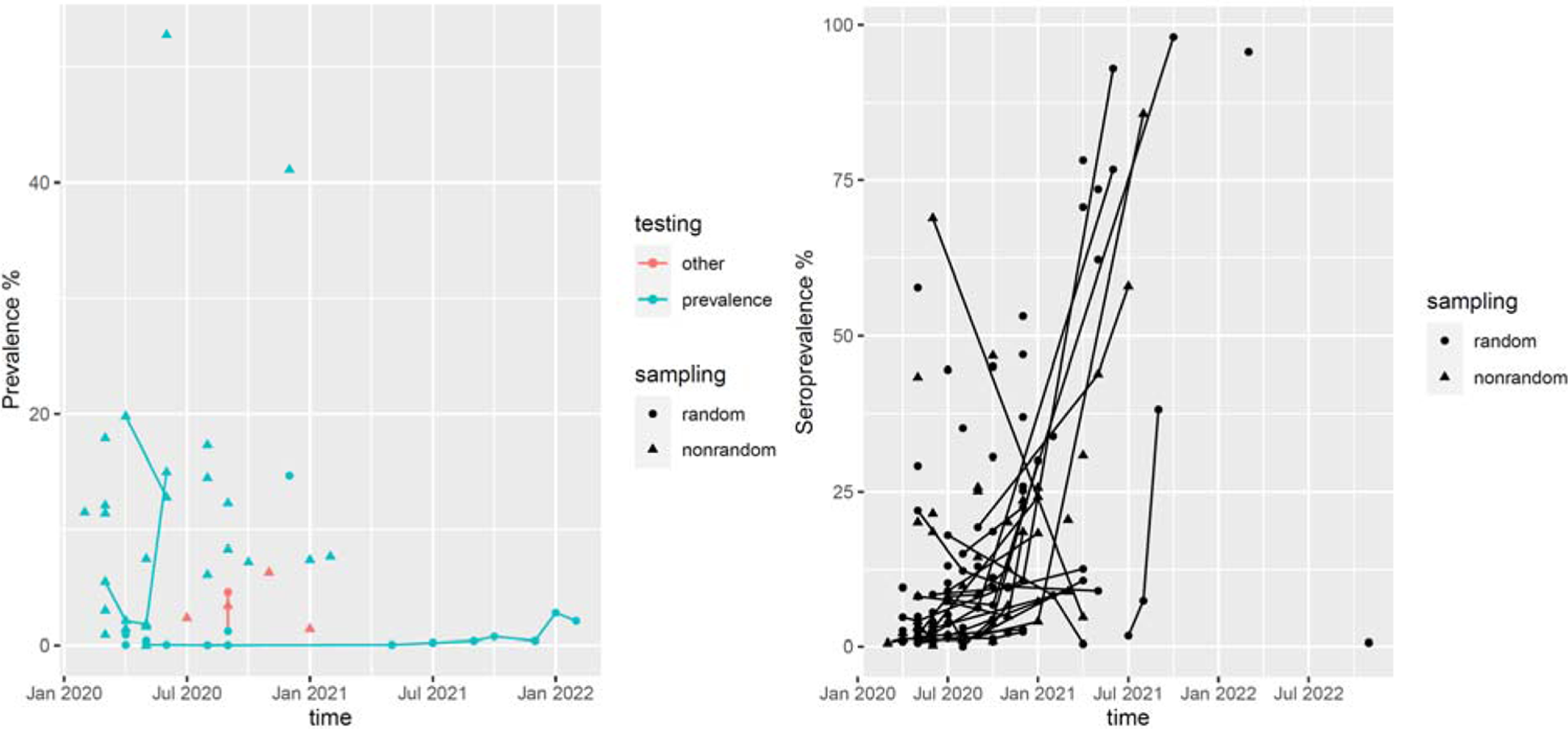
Estimated prevalence (left) and seroprevalence (right) by month, sampling, and testing methods.

**Figure 3.**
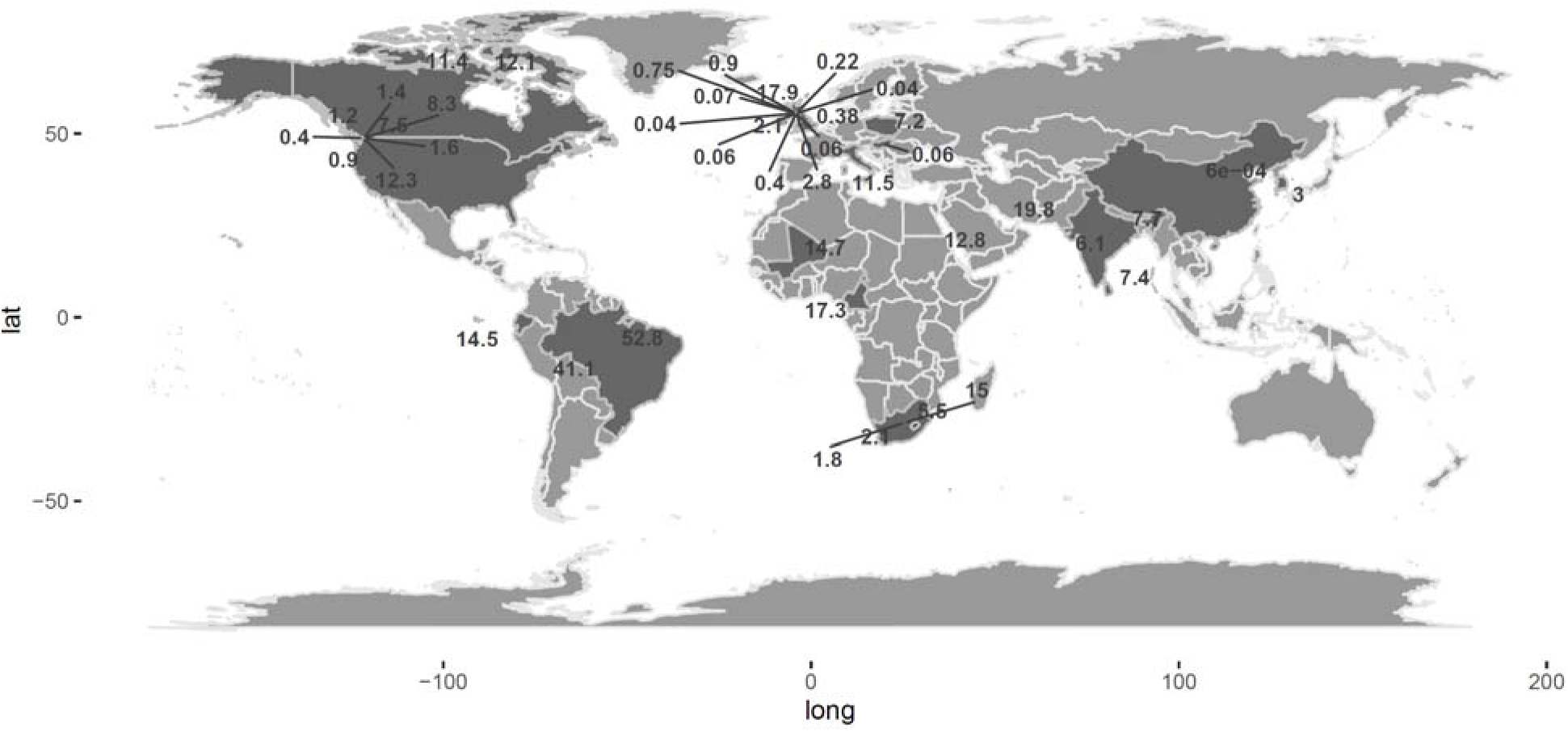
Map of COVID-19 prevalence in a global view.

**Figure 4.**
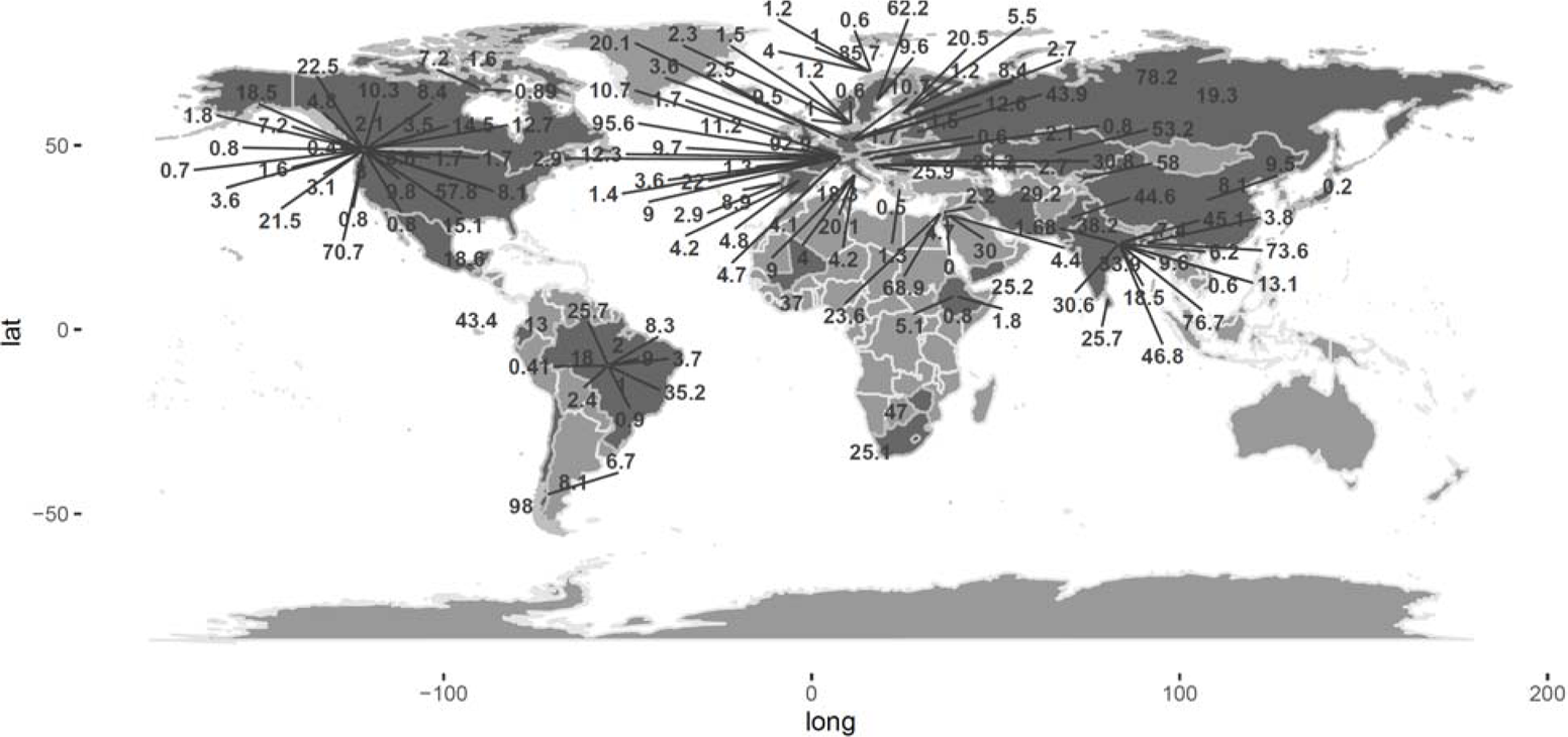
Map of COVID-19 seroprevalence in a global view.

Nineteen studies measured prevalence of COVID-19 only, 95 studies measured seroprevalence (i.e., antibody level) only, and 9 studies measured both prevalence and seroprevalence. Three studies^32–34^ used a mixture of PCR, antibody, and/or self-reported COVID-19 history. For the 28 studies reporting COVID-19 prevalence, 26 studies were based on RT-PCR tests (on swab or blood samples, or both), one study was based on an antigen test^35^, and one study was based on both antigen and RT-PCR tests^36^. For studies of seroprevalence, most studies used testing methods targeting on either IgA, IgM, or IgG. Two studies used anti-RBD antibodies^37,38^, and two studies changed targeting antibody in the middle of the sample collection^34,39^.

### Prevalence studies and comparison with younger age group and WHO reports

In total, 28 studies in 16 countries reported estimates of prevalence, either based on RT-PCR or antigen tests. All studies were conducted in 2020 or 2021, except for REACT-1 where the most recent round published was conducted in February and March 2022^40^. The prevalence of COVID-19 ranged diversely, from 0.006%^41^ in China to 52.8%^42^ in Brazil in 2020, and from 0.04%^43^ in the UK to 41.1%^44^ in Brazil in 2021. Study 30^40,43,45–47^ and Study 89^48^ had multiple rounds of sample collection. For Study 30 in England, the prevalence measurement started from May 2020, decreased in July – September 2020, increased thereafter until January 2022, and decreased in the latest round available in March 2022 (round 18). For Study 89 in South Africa, four rounds of sample collection were conducted monthly from March 2020. Prevalence decreased in April and May 2020 and increased dramatically (from 1.8% to 15%) in June 2020.

The comparison with prevalence in younger age groups is represented in Table 3 and Figure 5. 32% studies (9/28, 12/42 estimates) found a higher infection prevalence among older people compared with younger people. Four studies estimated significantly higher prevalence among the older people, while 6 studies estimated significantly lower prevalence.

**Figure 5.**
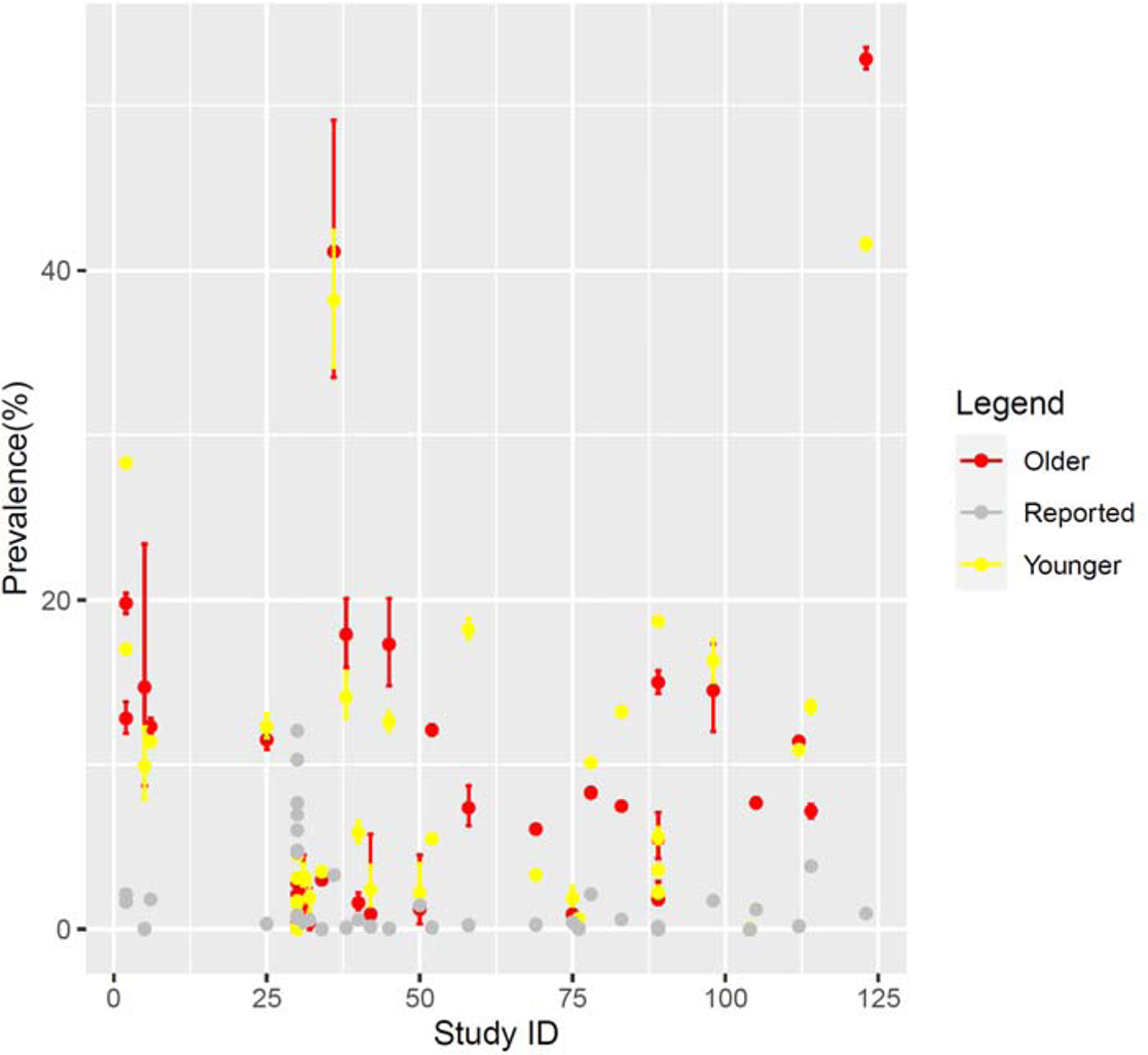
Comparison between study-based estimates and surveillance-based estimates.

**Table 3.** Comparison between study-based estimates and surveillance-based estimates.

| ID | Country | Estimates % (A) | WHO Reported % (B) | Ratio (B/A) | Younger group % (C) | Ratio (C/A) |
| --- | --- | --- | --- | --- | --- | --- |
| 2 | Qatar | 19.8 (19.2, 20.4) | 1.65 | 0.13 | 28.3 (28.2, 28.4) | 1.33 |
| 2 | Qatar | 12.8 (11.9, 13.8) | 2.13 | 0.11 | 17.0 (16.7, 17.2) | 1.43 |
| 5 | Mali* | 14.7 (8.7, 23.4) | 0.03 | <0.01 | 9.9 (7.9, 12.3) | 0.67 |
| 6 | USA | 12.3 (11.8, 12.8) | 1.81 | 0.15 | 11.4 (11.3, 11.5) | 0.93 |
| 25 | Italy | 11.5 (10.9, 12.1) | 0.34 | 0.03 | 12.3 (11.6, 13.1) | 1.07 |
| 30 | England | 0.07 (0.05, 0.12) | 0.65 | 9.33 | 0.15 (0.12, 0.17) | 2.14 |
| 30 | England | 0.06 (0.04, 0.09) | 0.79 | 13.15 | 0.08 (0.07, 0.10) | 1.33 |
| 30 | England | 0.04 (0.02, 0.06) | 0.82 | 20.48 | 0.03 (0.02, 0.04) | 0.75 |
| 30 | England | 0.04 (0.03, 0.07) | 0.85 | 21.28 | 0.11 (0.09, 0.13) | 2.75 |
| 30 | England | 0.06 (0.04, 0.1) | 4.65 | 77.50 | 0.15 (0.12, 0.18) | 2.50 |
| 30 | England | 0.22 (0.18, 0.29) | 4.80 | 21.81 | 0.66 (0.60, 0.73) | 3.00 |
| 30 | England | 0.38 (0.31, 0.45) | 6.04 | 15.90 | 0.92 (0.85, 0.99) | 2.42 |
| 30 | England | 0.75 (0.66, 0.86) | 6.98 | 9.31 | 1.7 (1.6, 1.8) | 2.27 |
| 30 | England | 0.40 (0.33, 0.48) | 7.71 | 19.27 | 1.6 (1.5, 1.7) | 4.00 |
| 30 | England | 2.8 (2.6, 3.0) | 10.30 | 3.68 | 4.5 (4.3, 4.6) | 2.14 |
| 30 | England | 2.1 (1.9, 2.3) | 12.05 | 5.74 | 3.1 (3.0, 3.2) | 1.11 |
| 31 | USA | 1.4 (0.4, 4.5) | 0.42 | 0.30 | 3.2 (2.6, 4.0) | 2.29 |
| 32 | USA | 0.4 (0.0, 2.5) | 0.55 | 1.39 | 1.9 (1.1, 3.2) | 4.75 |
| 34 | South Korea* | 3.0 (2.8, 3.1) | 0.01 | <0.01 | 3.5 (3.4, 3.6) | 1.17 |
| 36 | Brazil | 41.1 (33.5, 49.1) | 3.29 | 0.08 | 38.2 (34.1, 42.4) | 0.93 |
| 38 | UK | 17.9 (15.9, 20.1) | 0.11 | 0.01 | 14.1 (12.8, 15.6) | 0.79 |
| 40 | USA | 1.6 (1.2, 2.2) | 0.56 | 0.35 | 5.9 (5.3, 6.6) | 3.69 |
| 42 | England | 0.9 (0.0, 5.8) | 0.17 | 0.18 | 2.4 (1.4, 3.9) | 2.67 |
| 45 | Cameroon* | 17.3 (14.8, 20.1) | 0.05 | <0.01 | 12.6 (12.0, 13.2) | 0.73 |
| 50 | USA | 1.2 (0.3, 4.5) | 1.45 | 1.21 | 2.2 (1.2, 4.0) | 1.83 |
| 52 | Canada | 12.1 (11.8, 12.4) | 0.11 | 0.01 | 5.5 (5.3, 5.7) | 0.45 |
| 58 | Sri Lanka* | 7.4 (6.3, 8.7) | 0.25 | 0.03 | 18.2 (17.6, 18.9) | 2.46 |
| 69 | India* | 6.1 (5.9, 6.3) | 0.29 | 0.05 | 3.3 (3.2, 3.3) | 0.54 |
| 75 | USA | 0.9 (0.4, 1.5) | 0.42 | 0.46 | 1.9 (1.4, 2.6) | 2.11 |
| 76 | Hungary | 0.06 (0.0, 0.16) | 0.06 | 1.02 | 0.63 (0.47, 0.84) | 10.50 |
| 78 | USA | 8.3 (8.3, 8.4) | 2.12 | 0.26 | 10.1 (10.1, 10.1) | 1.22 |
| 83 | USA | 7.5 (7.3, 7.7) | 0.58 | 0.08 | 13.2 (13.1, 13.4) | 1.76 |
| 89 | South Africa* | 5.5 (4.3, 7.1) | 0.001 | <0.01 | 5.6 (5.2, 6.2) | 1.02 |
| 89 | South Africa* | 2.1 (1.5, 2.9) | 0.01 | <0.01 | 2.2 (2.0, 2.6) | 1.05 |
| 89 | South Africa* | 1.8 (1.5, 2.2) | 0.03 | 0.02 | 3.6 (3.4, 3.8) | 2.00 |
| 89 | South Africa* | 15.0 (14.3, 15.7) | 0.15 | 0.01 | 18.7 (18.5, 19.0) | 1.25 |
| 98 | Ecuador | 14.5 (12.0, 17.3) | 1.73 | 0.12 | 16.3 (15.0, 17.6) | 1.12 |
| 104 | China | 0.0006 (0.0000, 0.0040) | 0.000008 | 0.01 | 0.05 (0.03, 0.10) | 83.33 |
| 105 | India * | 7.7 (7.4, 8.0) | 1.19 | 0.15 | Not reported | - |
| 112 | Canada | 11.4 (11.2, 11.7) | 0.20 | 0.02 | 10.9 (10.7, 11.1) | 0.96 |
| 114 | Poland | 7.2 (6.8, 7.6) | 3.84 | 0.53 | 13.5 (13.1, 13.9) | 1.88 |
| 123 | Brazil | 52.8 (52.2, 53.5) | 0.95 | 0.02 | 41.6 (41.2, 41.9) | 0.79 |

Note: Age-specific cases were not available from WHO in countries with \*, so all-age prevalence was calculated instead.

The comparison between study-based estimates and WHO-reported estimates are as shown in Table 3 and Figure 5. For seven of the included prevalence studies^27,48–53^ (marked with * in Table 3), WHO reported daily cumulative COVID-19 cases were not available when stratified by age group, thus all-ages prevalence was used as a comparator. Out of the 28 studies, 4 studies from England ^40,43,45–47^, United States^54,55^, and Hungary^56^ reported a lower prevalence compared to the reported estimates (ratio of study-based estimates versus WHO reported estimates: 1.02 - 77.5). Twenty-three studies reported higher prevalence compared to the WHO reported estimates (ratio: <0.01 ∼ 0.53), and 14 studies (50%) reported at least ten times higher prevalence compared to WHO figures.

### Seroprevalence studies

In total, 104 studies reported estimates of seroprevalence. Twenty-seven studies were national studies whereas 66 studies were regional or sub-national studies. In 2020, seroprevalence was estimated ranging from 0% in Jordan^57^ to 22.5% in United States^58^, from 0.41% in Brazil^59^ to 98% in Chile^60^ in 2021.

Twenty-five studies reported multiple rounds of sample collection. The temporal trend of COVID-19 seroprevalence for 23 of the studies is shown in Figure 2. Two studies^61,62^ were excluded as they only reported adjusted estimates for subgroups.

## Discussion

This review covers prevalence estimates from 50 countries or regions from January 2020 to March 2022, but prevalence in these areas only represent a small part of the COVID-19 infection and vaccination among older adults in the world. Along with other reviews^63,64^, this scoping review shows that prevalence and seroprevalence studies were concentrated in several high-resource countries or areas such as the United States and Europe. Although our search included most of 2022, most included studies were conducted in 2020 and 2021; this may have reflected an urgent need to obtain prevalence estimates early on in the pandemic as well as the time it takes for the publishing process.

Twelve out of 42 estimates of prevalence were higher in older people, which indicates that the older adults were more likely to get infected in these areas. For the rest (30/42 estimates), a lower prevalence among older adults was observed comparing with younger groups in these regions during the sample collection time. Other primary studies and literature reviews also show a divergent age effect on infection. Zhang et al^65^ found that the infection rate was higher in older adults than people aged 15 to 64 years in February 2020 while Rumain et al^66^ found a higher prevalence among adolescents in summer 2020. Bergeri et al^10^ found a lower prevalence among people aged 0 to 9 years 10 to 19, and 60 and above across 2020-2022, comparing with young adults aged 20 to 29 years. A modeling study in Brazil^67^ showed little age effect in the difference of infection risks between regions. Nevertheless, studies have been agreed upon older age being a risk factor of worse outcomes of COVID-19 infection, including severe symptoms, hospitalization, and mortality^13–15^. Policies and measurements have been advocated and implemented to protect older people^68^ across the pandemic, however, the efficiency remains concerning in some countries.

Regardless of the age group, we found universal discrepancies between study-based prevalence and WHO reported prevalence where most of the studies reported a much higher prevalence. Besides underreporting, alternative reasons such as nonrepresentative population selection, lack of adjustment for test performance and study design, and delay of case reporting, may also lead to the difference. For example, participants in Study 104^41^ were individuals who were tested for COVID-19 in Zhongnan hospital of Wuhan University, and tests were only available for people with symptoms^69^, which may cause the prevalence in the study population to be significantly higher than the prevalence among general population. Nevertheless, infections estimated based on seroprevalence were consistently found to exceed the reported cases globally^9,10,70–72^. Monitoring current infection rate helps scientists and the public to monitor the spread of the disease and to prevent health services shortage due to outbreaks. Therefore, inaccurate estimation of the prevalence can be misleading to decision makers and the allocation of health services.

Our review also calls for attention on the importance of reporting participants’ age distribution in COVID-19 studies. Comparing with previous reviews^9,10^, fewer studies were included in our review due to lack of age information. This can also be seen from Bergeri et al’s review^10^ and Bobrovitz et al’s review^9^ where only 23% (117/513) and 21% (127/605) of the studies reported seroprevalence of the older adults.

This scoping review focuses on presenting prevalence and seroprevalence of COVID-19 among the older adults. However, this review does have limitations. This review only searched for English-language studies until October 2022, which may omit important and the most recent materials. Seventy-six studies had age thresholds different from the WHO threshold of 65 years (mostly 60 years), which may lead to biased estimates of the ratio between WHO-reported and study-based prevalence. For seven of the included prevalence studies, WHO-reported daily cumulative COVID-19 cases were not available when stratified by age group for comparison. Thus, for these studies we could not directly compare the study-based and reported prevalence for older people.

## Conclusions

The prevalence and seroprevalence of COVID-19 have been crucial indicators of COVID-19 management. This review describes these two indicators among one of the most vulnerable populations, the older adults. Prevalence of COVID-19 ranges from 0.0006% in 2020 in a Chinese study to 52.8% in mid 2020 in a Brazilian study, and seroprevalence ranges from 0 in August 2020 in a Jordan study to 98% in late 2021 in a Chile study. Prevalence among the older adults were higher in one third of the studies compared with the younger group, but in most countries, both far exceeded WHO reported prevalence.

## Data Availability

All data produced in the present study are available upon reasonable request to the authors.

